# Rare Genetic Mechanisms in Dyggve-Melchior-Clausen Disease

**DOI:** 10.64898/2026.10.22.26365648

**Authors:** Prince Jacob, Lingjuan Hong, Deepa Bhat, KN Gopika, Shushu Huang, Benjamin Kramer, K Mohandas Nair, Monkol Lek, Siddaramappa J Patil

## Abstract

Dyggve-Melchior-Clausen disease (DMC) is a rare skeletal dysplasia caused by biallelic variants in *DYM*. We investigated four affected individuals from three families with classical DMC features and non-diagnostic whole-exome sequencing. Reanalysis identified a homozygous deep intronic variant, *DYM:* c.193+132G>T, in Family 1 and a homozygous 331-bp AluYb8 insertion in exon 12 in Family 2. CRISPR prime editing and transcript analysis demonstrated that c.193+132G>T activates cryptic splice sites, resulting in 13-bp and 67-bp pseudoexon inclusion predicted to disrupt the reading frame. A missense variant of uncertain significance was identified in Family 3. These findings expand the mutational spectrum of *DYM* and highlight the value of investigating non-coding and structural variants in patients with a strong DMC phenotype.

## Main Text

Dyggve-Melchior-Clausen disease (DMC; OMIM #223800) is a rare autosomal recessive skeletal dysplasia characterized by disproportionate short stature, progressive spondylo-epi-metaphyseal abnormalities, microcephaly, and intellectual disability. Biallelic loss-of-function variants in *DYM*, encoding the Golgi-associated protein Dymeclin, cause DMC, whereas missense variants are more commonly associated with the allelic disorder Smith–McCort dysplasia, which lacks intellectual disability^1^. Deep intronic splice-altering variants and mobile element insertions (MEIs) are increasingly recognized as causes of rare diseases^2,3^. In *DYM*-associated DMC, however, both remain poorly characterized, and neither is reliably detected by routine diagnostic pipelines. Here, we investigated four affected individuals from three unrelated families with classical DMC and identified two novel disease mechanisms in *DYM*, including a deep intronic splice-altering variant, and an Alu-element insertion. Functional validation using CRISPR prime editing and transcript analysis demonstrated that the deep intronic variant activates cryptic splice sites and induces inclusion of pseudoexons, expanding the molecular mechanisms underlying *DYM*-associated DMC.

Four affected individuals (P1-P4) from three unrelated Indian families (F1, F2, F3) with classical clinical features of DMC were investigated (Figure 1A). Whole-exome sequencing (WES) was performed in one affected individual per family (P2, P3 and P4) and was non-diagnostic in each. Detailed methodology used in the study is provided in the Supplementary Methods.

**Figure 1.**
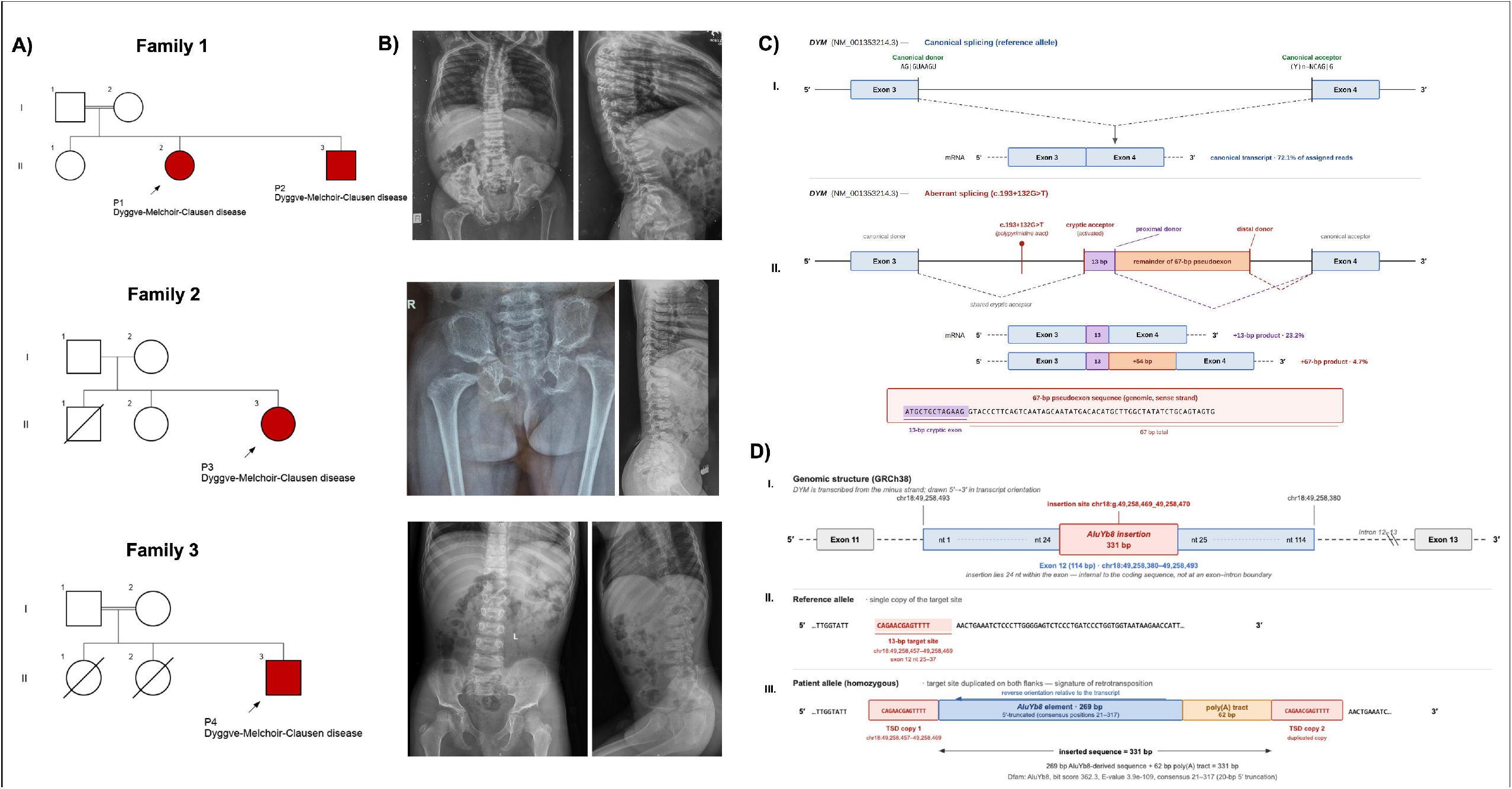
Clinical, molecular, and mechanistic characterization of *DYM* variants in Dyggve–Melchior–Clausen disease. (A) Pedigrees of the three unrelated families showing segregation of the DMC phenotype; affected individuals are indicated in red. (B) Representative skeletal radiographs of affected individuals from the corresponding families showing characteristic features of DMC. (C) Schematic representation of the effect of the deep intronic *DYM* variant c.193+132G>T on pre-mRNA splicing. In the reference allele, exon 3 is spliced directly to exon 4 using the canonical donor and acceptor sites. The variant lies within the polypyrimidine tract of a cryptic acceptor in intron 3 and activates it; this shared acceptor is used by two aberrant transcripts that differ in donor-site selection, generating a 13-bp inclusion (proximal donor) and a 67-bp pseudoexon (distal donor). Percentages indicate the fraction of assigned amplicon-sequencing read pairs from prime-edited HEK293T cells. Both aberrant products disrupt the reading frame and introduce a premature termination codon, predicting nonsense-mediated decay. (D) Genomic and sequence-level characterization of the *DYM* AluYb8 insertion. The homozygous 331-bp insertion lies 24 nt within the coding sequence of exon 12 (chr18:49,258,469_49,258,470; GRCh38) and comprises a 269-bp AluYb8 element in reverse orientation relative to the transcript and a 62-bp poly(A) tract. It is flanked by identical 13-bp target-site duplications (CAGAACGAGTTTT), consistent with retrotransposition-mediated insertion. The insertion disrupts the reading frame and introduces a premature termination codon. Sizes are not drawn to scale.

Family 1 (Figure 1A) comprised two affected siblings (P1 and P2), born to consanguineous parents, who presented with classical DMC characterized by disproportionate short stature, developmental delay, microcephaly and characteristic skeletal abnormalities (Figure 1B). Singleton exome sequencing in P2 identified a homozygous deep intronic variant in *DYM* (NM_001353214.3): c.193+132G>T (Supplementary Figure 1A). Segregation analysis confirmed that both parents were heterozygous carriers and that the affected sibling was also homozygous for the variant (Supplementary Figure 1A). The variant was absent from the gnomAD and GenomeIndia databases, while being identified only once in the heterozygous state in the All of Us dataset (allele frequency: 1×10□). SpliceAI (Δ score 0.43), Pangolin (0.43) and Human Splicing Finder consistently predicted activation of a cryptic acceptor splice site. Furthermore, the SpliceAI-10k calculator predicted inclusion of a 67-bp pseudoexon. Based on the available evidence, the variant was initially classified as a variant of uncertain significance (VUS) (ACMG/AMP criteria: PM2, PP1, PP4).

Patient-derived fibroblasts from P1 and P2 were not available for RNA analysis, precluding direct assessment of the variant’s splicing effect in patient cells. We therefore modeled the variant at the endogenous *DYM* locus in HEK293T cells to functionally assess its effect on splicing. To determine whether the deep intronic variant directly altered pre-mRNA splicing, the c.193+132G>T substitution was introduced into wild-type HEK293T cells using CRISPR prime editing with the PEmax system and designed pegRNAs (Supplementary Methods). Successful genome editing was confirmed by genomic PCR and Sanger sequencing. Single-cell cloning yielded a clone (DYM_Clonal_25) for *DYM*: c.193+132G>T, in which EditR analysis of the Sanger trace estimated 55% edited (T) and 42% reference (G) alleles (Supplementary Figure 2B), consistent with a single edited allele; no clone carrying the edit on both alleles was recovered. All transcript analyses were therefore performed on this clone, in which canonical and aberrant products are expected to co-occur. RNA isolated from edited and wild-type cells was analysed by RT-PCR (Supplementary Figures: 1C). Wild-type cells produced the expected 169-bp amplicon of *DYM* (NM_001353214.3), whereas edited cells generated two additional amplicons of 182 bp and 236 bp (Supplementary Figure 1C). Sanger sequencing of the aberrant RT-PCR product confirmed inclusion of the 67-bp pseudoexon, and an embedded 13-bp cryptic exon sequence (Figure 1C; Supplementary Figure 1C). Quantification of targeted amplicon next generation sequencing results independently confirmed the canonical wild type, +13-bp and +67-bp products, which accounted for 72.1%, 23.2% and 4.7% of assigned read pairs respectively (Supplementary Figure 1D). Both aberrant products are predicted to cause frameshifts and premature termination codons, consistent with a loss-of-function mechanism. As these transcripts are substrates for nonsense-mediated decay and no NMD inhibition was performed, their steady-state abundance underestimates the true extent of mis-splicing. These findings support reclassification of the variant to likely pathogenic (ACMG/AMP criteria: PS3, PM2, PP1, PP4).

Family 2 consisted of a girl (P3), who presented with classical clinical and radiological features of DMC (Figure 1B). Manual inspection of exome alignment in Integrative Genomics Viewer (IGV) identified clusters of soft-clipped reads at the exon 12–intron 11 junction of *DYM*, suggestive of structural alteration (Supplementary Figure 3A). BLAT search of the soft-clipped sequences indicated homology to an Alu/SINE sequence. PCR amplification followed by Sanger sequencing of the aberrant amplicon (Supplementary Figure 3B) confirmed a homozygous 331-bp Alu transposable mobile element insertion (MEI) within the coding sequence of exon 12 of *DYM* (NM_001353214.3). Dfam database analysis (Supplementary Figure 4) identified the insertion as belonging to the AluYb8 subfamily (mobile and active). The inserted sequence comprised a 269-bp AluYb8 element followed by a 62-bp poly(A) tract and was flanked by identical 13-bp target-site duplications, consistent with a retrotransposition-mediated insertion (Figure 1D). Insertion of the 331-bp MEI within the coding sequence is predicted to disrupt the reading frame and introduce a premature termination codon. The insertion was present in the heterozygous state in both parents and the unaffected sibling (Supplementary Figure 3B). These findings support identification of a pathogenic structural variant (ACMG/AMP criteria: PVS1, PM2, PP4).

Lastly, in Family 3, the affected boy (P4) had short stature, intellectual disability and characteristic skeletal abnormalities, platyspondyly, double-humped vertebrae, irregular vertebral endplates and lacy iliac crests (Figure 1B). Exome sequencing identified the homozygous variant, *DYM:* c.1460G>A (p.Ser487Asn), which was absent from aforementioned population databases, and parental segregation analysis confirmed that both parents were heterozygous carriers. The variant was classified as a VUS (ACMG/AMP criteria: PM2, PP3, PP4). Functional confirmation of p.Ser487Asn will require assessment of Dymeclin stability and subcellular localization, the approach previously used to establish pathogenicity for other DMC-associated missense variants^4^.

The two rare mechanisms identified here both predict complete loss of Dymeclin function, which raises the question of how variant type relates to phenotype in *DYM*-associated diseases. Dymeclin (DYM) is a Golgi-associated protein involved in intracellular vesicular trafficking, and Dymeclin deficiency has been shown to disrupt Golgi organization and vesicular trafficking, providing a cellular basis for the pathogenesis of DMC^4^. The genotype–phenotype spectrum of *DYM*-associated DMC may depend on the degree and nature of residual Dymeclin function. Most variants reported in DMC are predicted loss-of-function alleles, whereas *DYM* missense variants associated with Smith–McCort dysplasia have been proposed to retain residual Dymeclin activity, potentially accounting for the absence of intellectual disability in this phenotype^1^. However, missense variation is not invariably milder: the DMC-associated p.Asn469Tyr causes abnormal Dymeclin localization and aggregation^4^. It lies in the same conserved PF09742 domain as p.Ser487Asn identified here. Thus, the clinical consequence of *DYM* variation may reflect the residual activity, stability, or subcellular localization of Dymeclin rather than variant class alone.

Our findings further expand the mutational landscape of *DYM*-associated DMC. Deep intronic variants can activate otherwise cryptic splice sites, resulting in incorporation of intronic sequence as pseudoexons into mature transcripts, which can cause frameshifts and premature termination codons^2^. In our study, the c.193+132G>T variant was experimentally demonstrated to activate cryptic splice sites and cause inclusion of 13-bp and 67-bp pseudoexons via a shared cryptic acceptor and competing donor sites, providing direct functional evidence for its pathogenic effect. Notably, the variant lies within the polypyrimidine tract of the activated cryptic acceptor rather than within a splice-site motif and introduces a new pyrimidine; a configuration proposed to enhance U2AF65 recruitment and one that accounts for only a small minority of catalogued pseudoexon-instigating variants^5^.

The second mechanism, mobile element insertion, is mechanistically unrelated but converges on the same outcome. MEIs can disrupt coding sequences and normal gene function. Alu elements can cause disease through insertional mutagenesis and disruption of coding sequences or splicing, and MEIs may be missed by conventional short-read variant-calling pipelines^3^. The homozygous AluYb8 insertion identified here directly disrupts the *DYM* coding sequence and further expands the range of structural variation associated with DMC.

Together, these findings highlight the importance of considering such novel variations in unresolved patients with a strong DMC phenotype. Integration of multi-omic approaches can uncover pathogenic mechanisms that may otherwise remain undetected in Mendelian diseases.

## Data Availability

All data produced in the present study are available upon reasonable request to the authors

https://github.com/benjaminkramer510/dym-amplicon-analysis

## CRediT Author Contribution Statement

Prince Jacob: Data curation; Formal analysis; Funding acquisition; Investigation; Methodology; Visualization; Writing – original draft; Writing – review & editing. Lingjuan Hong: Formal analysis; Investigation; Methodology; Validation; Writing – review & editing. Deepa Bhat: Data curation; Formal analysis; Funding acquisition; Investigation; Writing – review & editing. Gopika K. N.: Data curation; Investigation; Writing – review & editing. Shushu Huang: Formal analysis; Validation; Writing – review & editing. Benjamin Kramer: Formal analysis; Software; Validation; Writing – review & editing. Mohandas Nair K: Data curation; Investigation; Writing – review & editing. Monkol Lek: Conceptualization; Funding acquisition; Methodology; Supervision; Writing – review & editing. Siddaramappa J. Patil: Conceptualization; Funding acquisition; Methodology; Supervision; Writing – review & editing.

## Ethics Declaration

The Institutional Review Board (IRB) of the Public Health Research Institute of India, Mysuru, India (approval no. 2025-09-01-97), the Institutional Ethics Committee of JSS Medical College, Mysuru, India (approval no. JSSMC/IEC/972025/13 NCT/2024-25), and IRB of Yale University (ID: 2000041040) gave ethical approval for this work. Written informed consent was obtained from the participants and/or their legal guardians for participation in the study and for the use of clinical and genetic information for research and publication.

## Data availability

The data used in the study is available from the corresponding author upon reasonable request.

## Acknowledgements

We thank the patients and their families for participating in this study and for providing clinical information and biological samples that enabled this work. We thank Dr Purnima Madhivanan (Mel and Enid Zuckerman College of Public Health, University of Arizona, Tucson, Arizona, USA), Dr Kiranmayee Muralidhar and Mrs Poornima Jayakrishna (Public Health Research Institute of India, Mysuru, India) for their guidance and institutional support. The PEmax expression construct in a Sleeping Beauty transposon backbone was a gift from Dr Evgueni (Zhenya) Ivakine, The Hospital for Sick Children, Toronto, Canada.

