## Supplementary Methods for "Rare Genetic Mechanisms in Dyggve-Melchior-Clausen Disease"

**B1. CRISPR Prime Editing and Transcript-Level Functional Validation of the *DYM* c.193+132G>T Deep Intronic Variant**

### Experimental overview

The deep intronic DYM variant c.193+132G>T (NM_001353214.3; GRCh38 chr18:49,391,461C>A) was evaluated by CRISPR prime editing in HEK293T cells. The experimental workflow comprised design and construction of engineered prime editing guide RNAs (epegRNAs), generation of a PEmax-expressing HEK293T cell population, introduction of the variant at the endogenous *DYM* locus, genomic confirmation of editing, and transcript-level assessment by RT-PCR and sequencing. Three primer binding site (PBS) and reverse transcription template (RTT) configurations were evaluated, and the configuration showing the highest observed editing efficiency was selected for downstream analyses.

### 1. Cell line and prime-editing system

HEK293T cells were used for functional analysis of the endogenous DYM locus and were maintained in Dulbecco's modified Eagle medium (DMEM) supplemented with 10% fetal bovine serum and 1× Antibiotic-Antimycotic solution (Gibco, Thermo Fisher Scientific, Waltham, MA, USA; cat. no. 15240-062) at 37°C in a humidified incubator with 5% CO₂. The prime-editing system consisted of PEmax and a puromycin-selectable lenti-puro-epegRNA construct.

| **Parameter** | **Final experimental setting** |
| --- | --- |
| Cell line | HEK293T |
| Prime editor | PEmax |
| epegRNA backbone | lenti-puro-epegRNA |
| Target | NM_001353214.3: c.193+132G>T |
| Genomic coordinate | GRCh38 chr18:49,391,461C>A |

### 2. Generation of a stable PEmax-expressing HEK293T cell population

To generate a stable PEmax-expressing cell population, HEK293T cells were co-transfected with a Sleeping Beauty transposon donor plasmid encoding PEmax with a blasticidin-resistance marker together with an SB100X transposase expression plasmid. The PEmax donor and SB100X plasmids were transfected at a plasmid mass ratio of 1:2 using Lipofectamine 3000 according to the manufacturer's instructions. At 24 h after transfection, cells were subjected to selection with 5 µg/mL blasticidin (prepared from a 10 mg/mL stock solution; Gibco, Thermo Fisher Scientific, Waltham, MA, USA; cat. no. A11139-03) for 7 days. Selection was continued until the non-transfected control population was eliminated. Surviving cells were expanded as a polyclonal HEK293T-PEmax population and maintained under blasticidin selection. PEmax activity was validated using a positive control epegRNA, *FKTN* (NM_001079802.2): c.139C>T, which showed an editing efficiency of approximately 24%, was included as a positive control).

### 3. epegRNA design

### Three epegRNAs targeting the *DYM* deep intronic variant c.193+132G>T were designed using pegFinder with default parameters (Chow et al., 2021). A 300-bp genomic sequence centered on the target nucleotide, together with the corresponding edited sequence (both provided below; designated by green-wildtype nucleotide and red-edited nucleotide), was used as input. pegFinder returned candidate spacer, PBS and RTT sequences. PBS lengths were selected around the recommended starting length of approximately 13 nt (Anzalone et al., 2019), consistent with high-throughput analyses reporting highest efficiency for PBS lengths of 11–13 nt (Kim et al., 2021). Longer reverse-transcription templates were required because the target nucleotide lies distal to the nick site; RT templates were designed to extend well beyond the edit to provide sufficient 3′ flap homology, in line with published guidance for long-range edits (Anzalone et al., 2019) and with analyses showing that extended RTT lengths substantially increase the range of targetable variants (Morris et al., 2021). On this basis, three PBS+RTT configurations were selected — epegRNA-A (PBS 13 + RTT 37), epegRNA-B (PBS 13 + RTT 38) and epegRNA-C (PBS 14 + RTT 37) — each combined with the same targeting spacer and scaffold (Supplementary Table S2). Candidate designs were evaluated experimentally, and the configuration showing the highest observed editing efficiency (epegRNA-A) was carried forward. Oligonucleotide sequences are given in Supplementary Table S2.

### Sequence alignment:

### Wildtype:GGAGGAAGCAACCATTTCAGTCTGCAGGTCATTAGGTAAGTGGGAAAAATGTGTGTGGGGTGTTGTTTTCAAATTTTGTTTTAGATTTTGATAGTATAATATGAACTGAATTAATGAAGTAATTACTCTTCTTTGATAGCAATAGTAATAATTTTTTCTATTTGTGGCATTCACAGATGCTGCTAGAAGGTACCCTTCAGTCAATAGCAATATGACACATGCTTGGCTATATCTGCAGTAGTGGTAATGGACTCTGCTAAGCCATAGACACTCTTTCGTCTTGTTCTTTTTGTGAATTTT

### Edited:GGAGGAAGCAACCATTTCAGTCTGCAGGTCATTAGGTAAGTGGGAAAAATGTGTGTGGGGTGTTGTTTTCAAATTTTGTTTTAGATTTTGATAGTATAATATGAACTGAATTAATGAAGTAATTACTCTTCTTTGATAGCAATAGTAATAATTTTTTCTATTTGTGTCATTCACAGATGCTGCTAGAAGGTACCCTTCAGTCAATAGCAATATGACACATGCTTGGCTATATCTGCAGTAGTGGTAATGGACTCTGCTAAGCCATAGACACTCTTTCGTCTTGTTCTTTTTGTGAATTTT

### 4. epegRNA assembly

Each epegRNA combination was assembled from three annealed modules: a spacer duplex, a scaffold duplex, and a PBS+RTT duplex. The modules were inserted into the BsmBI-containing lenti-puro-epegRNA backbone using a BsmBI/T4 DNA ligase Golden Gate-type assembly strategy.

#### 4.1 Annealing and phosphorylation of oligonucleotide pairs

Spacer, scaffold, and PBS+RTT oligonucleotide pairs were prepared separately. For each duplex, the following components were combined in a total reaction volume of 10 µL:

| **Component** | **Volume** |
| --- | --- |
| Top oligonucleotide (100 µM) | 1.0 µL |
| Bottom oligonucleotide (100 µM) | 1.0 µL |
| 10× T4 DNA ligase buffer | 1.0 µL |
| T4 polynucleotide kinase | 0.5 µL |
| Nuclease-free water | 6.5 µL |
| Total | 10.0 µL |

Reactions were incubated at 37°C for 30 min, followed by 95°C for 5 min. The reactions were cooled to 25°C at 0.1–0.2°C/s and held at 12°C. Each annealed duplex was diluted 1:200 in nuclease-free water before assembly.

#### 4.2 Golden Gate-type epegRNA assembly

A separate 20-µL assembly reaction was prepared for each PBS+RTT configuration using the undigested lenti-puro-epegRNA backbone and the three annealed duplexes. BsmBI-v2 and T4 DNA ligase were included in the same reaction, so that digestion of the backbone and ligation of the modules occurred concurrently during cycling.

| **Component** | **Amount** |
| --- | --- |
| Undigested lenti-puro-epegRNA backbone | 75 ng |
| Spacer duplex (1:200) | 1.5 µL |
| Scaffold duplex (1:200) | 1.5 µL |
| PBS+RTT duplex (1:200) | 1.5 µL |
| 10× T4 DNA ligase buffer | 2.0 µL |
| BsmBI-v2 | 0.5 µL |
| T4 DNA ligase | 1.0 µL |
| Nuclease-free water | To 20 µL |
| Total | 20 µL |

Assembly reactions were subjected to 30 cycles of 42°C for 1 min and 16°C for 1 min, followed by 60°C for 5 min, with a final hold at 4°C.

#### 4.3 Transformation and plasmid verification

Aliquots (2–5 µL) of each assembly reaction were transformed into competent Escherichia coli cells. For heat-shock transformation, cells were incubated on ice for 30 min, heat shocked at 42°C for 30–45 s, returned to ice for 2 min, supplemented with SOC medium, and allowed to recover for 1 h at 37°C shaking at 250 rpm. Recovered cells were plated on selective agar containing the appropriate antibiotic for the epegRNA backbone. Three colonies were screened per assembly. Plasmid DNA was prepared from selected colonies, and the spacer, scaffold, PBS, RTT, module junctions, and overall insert orientation were verified by Sanger sequencing.

### 5. Prime editing of the endogenous *DYM* locus

Stable HEK293T-PEmax cells were seeded in 24-well plates one day before transfection. Cells were transfected separately with plasmids encoding epegRNA-A, epegRNA-B, or epegRNA-C using Lipofectamine 3000 according to the manufacturer's instructions. At 24 h after transfection, doxycycline was added to a final concentration of 3 µg/mL to induce PEmax expression. Cells carrying the epegRNA plasmids were enriched with 1 µg/mL puromycin dihydrochloride (Gibco, Thermo Fisher Scientific, Waltham, MA, USA; cat. no. A11138-03) for 5 days. Doxycycline was maintained during the editing period. Cells were collected on day 6 after transfection for genomic analysis.

### 6. Genomic DNA extraction and confirmation of editing

Genomic DNA was isolated from edited cells using QuickExtract DNA Extraction Solution (LGC Biosearch Technologies; cat. no. QE09050) according to the manufacturer's instructions. The region surrounding c.193+132G>T was amplified using locus-specific primers (Supplementary Table S2), generating an approximately 383-bp amplicon. PCR products were analysed by Sanger sequencing to confirm introduction of the intended nucleotide substitution, and editing efficiencies were quantified using EditR online tool. Edited populations were then taken forward for single-cell cloning.

### 7. Single-cell cloning and screening

Following prime editing and puromycin enrichment, cells were dissociated and seeded as single cells in 96-well plates by limiting dilution. Approximately 60 individual colonies were identified and expanded. As colonies reached sufficient confluency, cells were transferred sequentially into larger culture vessels. A fraction of each clone was collected for genomic DNA extraction while the remaining cells were maintained in culture during genotyping. The DYM target region was amplified by PCR and initially evaluated by Sanger sequencing. Candidate clones were further assessed by targeted amplicon sequencing to determine the proportions of edited and reference alleles and to identify unintended nucleotide substitutions or insertions/deletions near the editing site. Clones carrying the intended c.193+132G>T substitution were expanded, cryopreserved, and used for downstream analyses. The clone selected for downstream experiments (DYM_Clonal_25) carried the c.193+132G>T substitution a single edited allele state, with EditR (calculated as mutant signal divided by the sum of mutant and wild-type signals) estimating 55% edited (T) and 42% reference (G) signal at the target position (Supplementary Figure 2B); no homozygous clone was recovered.

### 8. RNA isolation, cDNA synthesis and RT-PCR

Total RNA was isolated from edited (DYM_Clonal_25) and unedited wild-type HEK293T cells using the NucleoSpin RNA kit (Macherey-Nagel, Düren, Germany; Cat. No. 740955.250) according to the manufacturer's instructions. First-strand cDNA was synthesized using the SuperScript III First-Strand Synthesis System for RT-PCR (Invitrogen, Thermo Fisher Scientific, Waltham, MA, USA; cat. no. 18080-051). The resulting cDNA was used as a template for RT-PCR amplification across the DYM exon 3–exon 4 region using locus-specific primers. Primers DYM_RTPCR_F and DYM_RTPCR_R (Supplementary Table S2) generated the expected 169-bp canonical product in wild-type cells, whereas edited cells additionally produced a 182-bp product, consistent with inclusion of the 13-bp cryptic exon, and a 236-bp product, consistent with inclusion of the 67-bp pseudoexon (Supplementary Figure 2).

### 9. Sanger sequencing and characterization of aberrant transcripts

RT-PCR products were separated by agarose gel electrophoresis. Individual bands were excised, purified, and analysed by Sanger sequencing to determine their exon composition and identify aberrant splice junctions. The 67-bp sequence identified in the aberrant transcript was:

**ATGCTGCTAGAAGGTACCCTTCAGTCAATAGCAATATGACACATGCTTGGCTATATCTGCAGTAGTG**

Within this sequence, the 13-bp cryptic exon sequence ATGCTGCTAGAAG is followed by the newly activated GT donor sequence. The longer 67-bp pseudoexon-containing transcript and the shorter 13-bp aberrant product therefore result from use of alternative cryptic donor sites. Inclusion of either intronic sequence is predicted to disrupt the coding reading frame and introduce a premature termination codon.

### 10. Targeted Next Generation Sequencing (NGS) of amplicon products

Targeted NGS of amplicon products using primers DYM_ampF and DYM_ampR (Supplementary Figure 2A) were additionally used to assess amplicon products generated following editing. The sequencing strategy distinguished the canonical exon 3–exon 4 junction from transcripts containing the predicted aberrant splice products. Reads supporting the canonical junction and the expected +13-bp and +67-bp products were identified.

| **Amplicons** | **Expected size** |
| --- | --- |
| Canonical product | 473 bp* |
| +13-bp product | 486 bp* |
| +67-bp product | 540 bp* |

*These sizes correspond to the DYM_ampF/DYM_ampR primer pair used for targeted amplicon sequencing. The shorter primer set used for the final RT-PCR assay generated the 169-bp canonical and 236-bp +67-bp products. The primer sets are mentioned in Supplementary Table S1.

**Data processing and analysis**

The pipeline for processing the targeted NGS amplicons is mentioned in the GitHub link in the Supplementary Table S3. For analysis, paired 300-bp reads were analyzed and checked for proper pairing, summarized read length, base quality, primer matching, ambiguous bases, and repeated sequence pairs. Splice products were classified using a 50-bp region of R2 that spanned the pseudoexon-exon 4 junction. There were three criteria for accepting a call. These include the best reference differing by no more than two bases, the second-best reference having at least four additional mismatches, and the R2 primer prefix having no more than two mismatches. The +67-bp calls were checked using an exact R1 junction sequence. The library lacked molecular identifiers, so reads were not deduplicated.

**B2. Whole exome sequencing (WES) and Data analysis**

WES was performed for P2, P3 and P4 using the capture kits outlined in the table below at Centre for DNA Finger Printing and Diagnostics, Hyderabad, India at Centre for DNA Finger Printing and Diagnostics, Hyderabad, India. For P2, target enrichment was performed using the Clinical Exome V8 + ncv capture kit with sequencing on an Illumina platform to a mean coverage of >80–100x. For P3 and P4, target enrichment was performed using the Nextera Rapid Capture Exome Kit with paired-end 2×150 bp sequencing on an Illumina NextSeq500 platform, achieving >95% of the target region covered at a mean coverage of >80–100x.

For all the samples, reads were aligned to the human reference genome (GRCh38.p13) using the Sentieon aligner, and alignments were subjected to duplicate removal, base quality score recalibration and indel realignment using Sentieon, following the GATK best practices framework for germline SNV and indel discovery from exome sequences. Variants were called using the Sentieon Haplotype Caller (v201808.07). Annotation was performed using VEP against the Ensembl release 99 human gene model and copy number variants were called from the targeted sequence data using ExomeDepth (v1.1.10). Variants were annotated against ClinVar, OMIM, GWAS, HGMD (v2020.2) and SwissVar, and filtered on allele frequency in 1000 Genomes Phase 3, gnomAD (v4.1.1), EVS, dbSNP (v151), and the 1000 Japanese Genome dataset.

| **Sequencing platform** | **Type of reads** | **Capture kit** | **Coverage** | **Individuals** |
| --- | --- | --- | --- | --- |
| NextSeq500 | PE 2×150 | Nextera Rapid Capture Exome Kit | >80-100x | P3, P4 |
| Illumina | PE 2×150 | Clinical Exome V8 + ncv | >80–100x | P2 |

**WES reanalysis**

Reanalysis of the primary sequencing data was performed using the Franklin platform (QIAGEN; formerly Genoox Franklin), applying its built-in germline exome analysis pipeline against GRCh38. In contrast to the standard clinical analysis, filtering was configured to retain intronic and other non-coding variants. Filters were applied on patient phenotype (HPO terms), genomic region, variant effect, zygosity, population frequency, and call confidence. The filtered variants were reviewed manually. In parallel, alignments were inspected in the Integrative Genomics Viewer (IGV) to identify structural signatures, such as clusters of soft-clipped reads, that are not reported by standard variant-calling pipelines.

#### B3. Genomic DNA extraction, PCR cycling conditions and Sanger sequencing

Genomic DNA was extracted from peripheral blood collected in EDTA from all affected individuals and available family members using the QIAamp DNA Blood Mini Kit (Qiagen, Hilden, Germany; cat. no. 51104) according to the manufacturer's instructions.

All endpoint PCR reactions described below were performed on an Applied Biosystems VeritiPro thermal cycler (Thermo Fisher Scientific, Waltham, MA, USA). Cycling conditions for each assay are given in Supplementary Table S1. PCR products and gel-excised bands were purified using the QIAquick PCR & Gel Cleanup Kit (Qiagen; Cat. No. 28506) according to the manufacturer's instructions before Sanger sequencing. The same extraction, purification, and instrumentation were used for all individuals in the study. DNA concentration and purity were measured using DeNovix DS-11 Spectrophotometer (DeNovix Inc., Wilmington, DE, USA).

#### Sanger sequencing and segregation analysis- Family 1

The region containing *DYM*: c.193+132G>T was amplified from genomic DNA by locus-specific PCR using primers DYM_gDNA_IF and DYM_gDNA_IR (Supplementary Table S2), generating an approximately 383-bp amplicon. Reactions were performed at a 50 μL volume, under the cycling conditions given in Supplementary Table S1. Products were resolved by agarose gel electrophoresis, purified using the QIAquick PCR & Gel Cleanup Kit and sequenced bidirectionally by Sanger sequencing. Chromatograms were visualised and analysed using Chromas software v2.6.6 (Technelysium Pty Ltd). The results are provided in Supplementary Figure 1A.

#### PCR and Sanger sequencing of the Alu element insertion – Family 2

The region spanning the insertion site in *DYM* exon 12 was amplified from genomic DNA using primers (DYM_I12F and DYM_I12R) flanking the insertion site (Supplementary Table S2). In the reference allele, this primer pair generates an approximately 679-bp product; in the presence of the 331-bp insertion, the product is approximately 1010 bp. Cycling conditions are provided in Supplementary Table S1. Products were resolved by agarose gel electrophoresis (Supplementary Figure 3B); both alleles were excised, purified using the QIAquick PCR & Gel Cleanup Kit and sequenced bidirectionally, and chromatograms were analysed using Chromas. The same primer pair was used to genotype both parents and the unaffected sibling. The schematic diagram is provided in Figure 1D.

#### Dfam database analysis of the AluYb8 inserted sequence

The 331-bp inserted sequence obtained by Sanger sequencing was submitted to the Dfam database (release 3.9) (Storer J, et al., 2021) through the web search interface at https://dfam.org on 29 October 2025 and searched using the default parameters. A single match was returned to the AluYb8 family (bit score 362.3, E-value 3.9e-109), spanning consensus positions 21 to 317. The web interface output is provided in Supplementary Figure 4.

#### PCR and Sanger sequencing of the missense variant – Family 3

Segregation testing for *DYM*: c.1460G>A (p.Ser487Asn) in Family 3 was performed by a third-party institute. Exon 13 of DYM was amplified by PCR and the product sequenced by Sanger sequencing. Sequences were aligned to the reference transcript ENST00000675505.1 and analysed using Chromas software v2.6.6 (Technelysium Pty Ltd). to confirm the substitution and determine zygosity in the proband and both parents.

#### Supplementary Table S1. PCR cycling conditions.

All reactions were performed on an Applied Biosystems VeritiPro thermal cycler in a 50µL volume.

| **Steps** | **Family 1 — DYM c.193+132G>T** | **Family 2 — AluYb8 insertion** |
| --- | --- | --- |
| Primers | DYM_gDNA_IF / DYM_gDNA_IR | DYM_I12F / DYM_I12R |
| Product size | 383 bp | ~679 bp (reference) / ~1010 bp (insertion) |
| Initial denaturation | 95 °C, 5 min, 1 cycle | 95 °C, 5 min, 1 cycle |
| Denaturation | 98 °C, 10 s | 98 °C, 10 s |
| Annealing | 59 °C, 30 s | 61 °C, 30 s |
| Extension | 72 °C, 30 s | 72 °C, 60 s |
| Cycles | 30 | 30 |
| Final extension | 72 °C, 5 min, 1 cycle | 72 °C, 5 min, 1 cycle |
| Hold | 4 °C, ∞ | 4 °C, ∞ |
| Polymerase | EmeraldAmp GT PCR Master Mix (2X premix; Code No. RR310A, Takara Bio, Kusatsu, Shiga, Japan) | EmeraldAmp GT PCR Master Mix (2X premix; Code No. RR310A, Takara Bio, Kusatsu, Shiga, Japan) |

**Supplementary Table S2.** Oligonucleotides used for epegRNA construction, PCR of variants, RT-PCR and targeted amplicon next generation sequencing.

Sequences are reported 5′→3′ as supplied in the oligonucleotide file. Lower-case bases and cloning overhangs are retained.

| **Oligonucleotide** | **Module** | **Sequence (5′→3′)** | **Scale** |
| --- | --- | --- | --- |
| DYM_SPA_TOP | Spacer | caccgTCATATTGCTATTGACTGAAGTTTT | 25 nmol |
| DYM_SPA_BOT | Spacer | CTCTAAAACTTCAGTCAATAGCAATATGAc | 25 nmol |
| DYM_SCAFTOP | Scaffold | AGAGCTAGAAATAGCAAGTTAAAATAAGGCTAGTCCGTTATCAACTTGAAAAAGTGGCACCGAGTCG | 40 nmol |
| DYM_SCAFBOT | Scaffold | GCACCGACTCGGTGCCACTTTTTCAAGTTGATAACGGACTAGCCTTATTTTAACTTGCTATTTCTAG | 40 nmol |
| DYM_pRA_TOP | PBS 13 + RTT 37 (epegRNA-A) | GTGCTTGTGtCATTCACAGATGCTGCTAGAAGGTACCCTTCAGTCAATAGCAAT | 25 nmol |
| DYM_pRA_BOT | PBS 13 + RTT 37 (epegRNA-A) | cgcgATTGCTATTGACTGAAGGGTACCTTCTAGCAGCATCTGTGAATGaCACAA | 25 nmol |
| DYM_pRB_TOP | PBS 13 + RTT 38 (epegRNA-B) | GTGCTTTGTGtCATTCACAGATGCTGCTAGAAGGTACCCTTCAGTCAATAGCAAT | 25 nmol |
| DYM_pRB_BOT | PBS 13 + RTT 38 (epegRNA-B) | cgcgATTGCTATTGACTGAAGGGTACCTTCTAGCAGCATCTGTGAATGaCACAAA | 25 nmol |
| DYM_pRC_TOP | PBS 14 + RTT 37 (epegRNA-C) | GTGCTTGTGtCATTCACAGATGCTGCTAGAAGGTACCCTTCAGTCAATAGCAATA | 25 nmol |
| DYM_pRC_BOT | PBS 14 + RTT 37 C(epegRNA-C) | cgcgTATTGCTATTGACTGAAGGGTACCTTCTAGCAGCATCTGTGAATGaCACAA | 25 nmol |
| **Oligonucleotides used for locus specific PCR of Family 1 for *DYM* variant: c.193+132G>T** | | | |
| DYM_gDNA_IF | NA | TGTCTCTCCAGTAGTGAGTTGAAA | 25 nmol |
| DYM_gDNA_IR | NA | TGGTGAACATAGTCAATAACAGCA | 25 nmol |
| **Oligonucleotides used for PCR of the AluYb8 insertion in *DYM* exon 12 identified in Family 2** | | | |
| DYM_I12F | NA | gtgctgcttaactcaccttca | 25 nmol |
| DYM_I12R | NA | accagaactgaaatttgctccc | 25 nmol |
| **Oligonucleotides used for RT-PCR of the HEK293T cells** | | | |
| DYM_RTPCR_F | NA | CAGGCACGGAATCTATCTCTGA | 25 nmol |
| DYM_RTPCR_R | NA | AGTGCACCAAGATTTCCTGTTC | 25 nmol |
| **Oligonucleotides used for Transcript amplicon NGS of HEK293T edited cells** | | | |
| DYM_ampF | NA | GGAAATGCAGGTTTAAGGACAA | 25 nmol |
| DYM_ampR | NA | AGTCAGAACTGTAATTGCCAGGA | 25 nmol |
| ill_DYM_F | NA | TCCCTACACGACGCTCTTCCGATCTAAGTAGAGGGAAATGCAGGTTTAAGGACAA | 25 nmol |
| ill_DYM_R | NA | GTTCAGACGTGTGCTCTTCCGATCTAGTCAGAACTGTAATTGCCAGGA | 25 nmol |

*NA: Not Applicable*

#### Supplementary Table S3. Software and in silico prediction tools

| Tools | Purpose | Link | Reference |
| --- | --- | --- | --- |
| SpliceAI (SpliceAI Lookup web interface, Broad Institute) | Splice-altering prediction | <https://spliceailookup.broadinstitute.org/> | 8 |
| SpliceAI-10k calculator (SAI-10k-calc) accessed via SpliceAI lookup web interface | Prediction of pseudoexon inclusion and resulting transcript |  | 4 |
| Pangolin (accessed via SpliceAI Lookup web interface) | Splice-altering prediction |  | 18 |
| Human Splicing Finder (HSF Pro / HSF 3.1) | Splice-site motif prediction | <https://genomnis.com/hsf> | 6 |
| Integrative Genomics Viewer (IGV) - v2.19.7 | Manual inspection of exome alignments; identification of soft-clipped reads | <https://igv.org/> | 14 |
| BLAT (UCSC) | Sequence identity search of soft-clipped reads | <https://genome.ucsc.edu/index.html> | 9 |
| Dfam/nhmmer (Release 3.9) | Repeat family assignment of the inserted sequence | <https://www.dfam.org/home> | 15 |
| pegFinder (web interface) | epegRNA design | <http://pegfinder.sidichenlab.org/> | 5 |
| EditR (web interface) | Quantification of prime editing efficiency from Sanger traces | <http://baseeditr.com/> | 11 |
| Chromas | For visualization of Sanger sequencing chromatograms | <https://technelysium.com.au/wp/chromas/> | NA |
| Amplicon Next Generation Sequencing analysis pipeline | Read assignment to canonical, +13-bp and +67-bp splice products; quantification | Code available at <https://github.com/benjaminkramer510/dym-amplicon-analysis> | NA |

#### *NA: Not Available*

#### B4. Databases and population frequency assessment

Allele frequencies were assessed in gnomAD (v4.1.1), the GenomeIndia Genome Variation Browser and the All of Us Research Hub Data Browser. GenomeIndia and All of Us were queried through their public web portals using GRCh38 coordinates (chr18:49,391,461 C>A for c.193+132G>T; chr18:49257010 C>T for *DYM*: c.1460G>A on [Access date: 1/24/26]. Both resources were interrogated for the specific alternate allele at each position.

The c.193+132G>T variant was absent from gnomAD v4.1.1, gnomAD SVs v4.1.0 and GenomeIndia databases and was observed in a single heterozygous individual in All of Us database (allele frequency 1×10⁻⁶). The c.1460G>A variant was absent from all three databases.

The AluYb8 insertion was not present in gnomAD SVs v4.1.0, and All of Us databases.

| **Database** | **Link** | **Reference** |
| --- | --- | --- |
| gnomAD (v4.1.1) | <https://gnomad.broadinstitute.org/> | 7 |
| GenomeIndia Genome Variation Browser | <https://gidb.igib.res.in/index.php> | 16 |
| All of Us Research Hub Data Browser | <https://databrowser.researchallofus.org/> | 17 |

#### B5. Variant classification

Variants were classified according to the ACMG/AMP standards and guidelines (Richards et al., 2015), incorporating the ClinGen Sequence Variant Interpretation Working Group recommendations for PVS1 (Abou Tayoun et al., 2018) and for functional evidence (Brnich et al., 2019). Criterion-by-criterion justification for each variant is provided in Supplementary Table S4.

### Supplementary Table S4. Justification of ACMG/AMP criteria applied to the three *DYM* variants identified in the study

Criteria were applied following Richards et al. (2015), with ClinGen SVI recommendations for PVS1 (Abou Tayoun et al., 2018) and for functional evidence (PS3/BS3; Brnich et al., 2020).

#### Variant 1 — *DYM* (NM_001353214.3): c.193+132G>T, homozygous (Family 1: P1, P2)

**Final classification: Likely pathogenic**

| Criterion | Strength applied | Evidence and justification |
| --- | --- | --- |
| PS3 | Strong | Prime editing of the variant into the endogenous *DYM* locus in HEK293T cells produced aberrant transcripts absent from isogenic unedited cells. RT-PCR gave an additional 236-bp product; Sanger sequencing confirmed inclusion of a 67-bp pseudoexon with an embedded 13-bp cryptic exon. Targeted amplicon NGS quantified +13-bp and +67-bp products at 23.2% and 4.7% of assigned read pairs, absent in wild-type. Assay validity: isogenic comparison, single-variant introduction, clinically relevant transcript readout, wild-type control included.  Caveat: One clone (single edited allele) was analysed and NMD inhibition was not performed. |
| PM2 | Moderate | Absent from gnomAD v4.1.1 and GenomeIndia databases; observed once in the heterozygous state in All of Us database (allele frequency 1×10⁻⁶). Consistent with a rare autosomal recessive disorder. Versions and query details in Supplementary Methods – B5. |
| PP1 | Supporting | Segregation with disease in Family 1: both affected siblings homozygous, both unaffected parents heterozygous. Applied at supporting only, as the number of informative meioses does not reach the threshold for Moderate or Strong. |
| PP4 | Supporting | Highly specific phenotype; classical clinical and radiological DMC including lacy iliac crests and double-humped vertebral bodies in a gene with a single established disease association. |

#### Variant 2 — *DYM* (NM_001353214.3) AluYb8 insertion in exon 12, homozygous (Family 2: P3)

**Final classification: Pathogenic**

| Criterion | Strength applied | Evidence and justification |
| --- | --- | --- |
| PVS1 | Very Strong | A 331-bp insertion within the coding sequence of exon 12 (chr18:49,258,469_49,258,470; GRCh38), predicted to disrupt the reading frame and introduce a premature termination codon. Per the ClinGen SVI PVS1 decision tree: (i) the variant is predicted to trigger NMD. (ii) *DYM* loss of function is the established disease mechanism for DMC; (iii) exon 12 is present in the MANE Select transcript and is not subject to appreciable alternative splicing. |
| PM2 | Moderate | Not present in gnomAD SVv4.1.0, and All of Us databases. |
| PP4 | Supporting | Classical clinical and radiological DMC phenotype in a gene with a single disease association. |

#### Variant 3 — *DYM* (NM_001353214.3): c.1460G>A (p.Ser487Asn), homozygous (Family 3: P4)

**Final classification: Variant of uncertain significance**

| Criterion | Strength applied | Evidence and justification |
| --- | --- | --- |
| PM2 | Moderate | Absent from gnomADv4.1.1, GenomeIndia and All of Us databases. |
| PP3 | Supporting | Concordant in silico prediction of a deleterious effect across missense predictors; the residue lies within the conserved Dymeclin (PF09742) domain. Applied at supporting per SVI recommendations for computational evidence. |
| PP4 | Supporting | Classical clinical and radiological DMC phenotype. |

**Abbreviations:** ACMG/AMP, American College of Medical Genetics and Genomics / Association for Molecular Pathology; NMD, nonsense-mediated decay; SVI, Sequence Variant Interpretation Working Group.

**Supplementary Figure Legends:**

**Supplementary Figure 1.** Functional validation of the *DYM* c.193+132G>T deep intronic variant and characterization of aberrant splice products. (A) Sanger sequencing of the *DYM:* c.193+132G>T locus in the proband; P1 (III.2), affected brother; P2 (III.3), and both parents (I.5 and II.3), demonstrating the variant in the affected individuals and the corresponding parental alleles. (B) Integrative Genomics Viewer (IGV) visualization of the WES data of P2 demonstrating *DYM* locus and c.193+132G>T variant along with the predicted cryptic splice sites within intron 3. The schematic indicates activation of the cryptic acceptor site and two alternative donor sites, resulting in inclusion of a shorter 13-bp cryptic sequence (corresponding to Band B indicated by green arrow in the gel image) and a 67-bp pseudoexon (corresponding to Band C indicated by blue arrow in the gel image) in the mature transcript. (C) RT-PCR analysis of *DYM* transcripts from wild-type and edited HEK293T cells showing the canonical transcript and aberrantly spliced products. The canonical transcript is 169 bp, whereas the larger products are 182-bp and 236-bp, consistent with inclusion of the 13-bp and 67-bp pseudoexons. Non-specific band of ~280-bp is observed in both the wildtype and edited cells. (D) Targeted amplicon-sequencing analysis of *DYM* transcript products. (a) Of 514,428 *DYM* amplicon read pairs, 460,425 were assigned to a splice-product category, comprising 331,993 reads supporting the canonical exon 3–exon 4 junction, 106,631 supporting the +13-bp aberrant product, and 21,801 supporting the +67-bp pseudoexon-containing product. (b) Of the 21,801 +67-bp calls identified using the downstream R2 junction, 20,333 were independently supported by an exact upstream +67-bp junction signature in R1, with no conflicting WT or +13-bp R1 signatures. (c) The fraction of +67-bp transcripts remained highly consistent across four classification rules, ranging from 4.728% to 4.752%.

**Supplementary Figure 2.** Amplicon design and confirmation of the prime-edited clone used for transcript analysis. (A) Primer positions and expected product sizes across the *DYM* exon 3–exon 4 junction. (I) Binding positions of the two primer pairs on the canonical *DYM* transcript (NM_001353214.3), drawn to scale. DYM_RTPCR_F and DYM_RTPCR_R amplify a 169-bp product spanning the exon 3–exon 4 junction, entirely nested within the 473-bp product amplified by DYM_ampF and DYM_ampR. The red line marks the exon 3–exon 4 junction, at which the pseudoexon is inserted in the aberrant transcripts. DYM_ampR spans the exon 5–exon 6 junction, so the amplicon-sequencing primer pair is cDNA-specific and does not amplify genomic DNA. (II) The three splice products includes: canonical transcript which joins exon 3 directly to exon 4; the +13-bp product which incorporates a 13-bp cryptic exon generated by use of a proximal cryptic donor site, and the +67-bp product incorporates the 67-bp pseudoexon generated by use of a distal cryptic donor site, with the 13-bp cryptic exon lying at the 5′ end of the 67-bp pseudoexon. Expected product sizes are given for both primer pairs, and both aberrant products are predicted to disrupt the reading frame. (B) Sanger chromatogram and EditR quantification of the prime-edited HEK293T clone (DYM_Clonal_25) used for all transcript analyses. The asterisk marks the edited position, at which the c.193+132G>T substitution is present at 55% alongside 42% of the reference G allele, consistent with a single edited allele. Base-calling percentages at each position are shown below the trace.

**Supplementary Figure 3.** Genomic validation and segregation of the *DYM* AluYb8 insertion in P3 (Family 2). (A) Integrative Genomics Viewer (IGV) visualization of the *DYM* locus in the proband (P3) showing reduced read coverage and soft-clipped reads within the coding sequence of exon 12, consistent with the presence of an insertion at the identified site. (B) PCR analysis of the region spanning the insertion site in the proband; P3 (III.3), unaffected sibling (III.2), father (II.3), mother (II.4), and an unrelated control. The proband shows the larger amplicon of approximately 1,000 bp corresponding to the 331-bp insertion, whereas the control shows the smaller approximately 679-bp reference amplicon. The unaffected sibling and both parents show both amplicons, consistent with heterozygosity for the insertion.

**Supplementary Figure 4.** Dfam database identification of the AluYb8 mobile element insertion in *DYM*. The inserted sequence (331 bp) identified in P3 (Family 2) was analysed using the Dfam transposable element database with the *Homo sapiens* reference set. The sequence showed a significant match to the AluYb8 family, with a bit score of 362.3 and an E-value of 3.9 × 10⁻¹⁰⁹.
