## Supplementary figures and images for "Rare Genetic Mechanisms in Dyggve-Melchior-Clausen Disease"

### Supplementary Figure 1

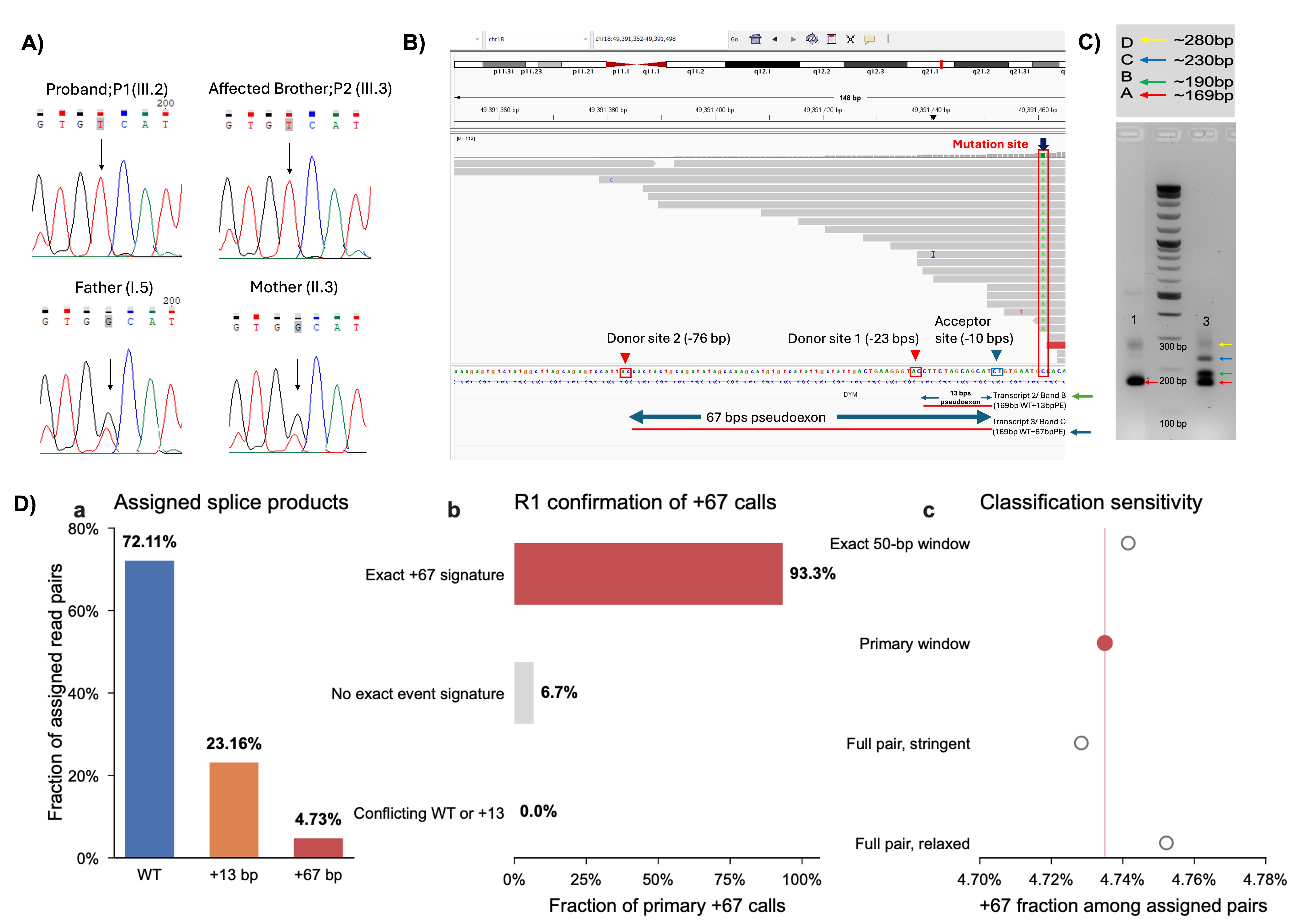

### Supplementary Figure 2

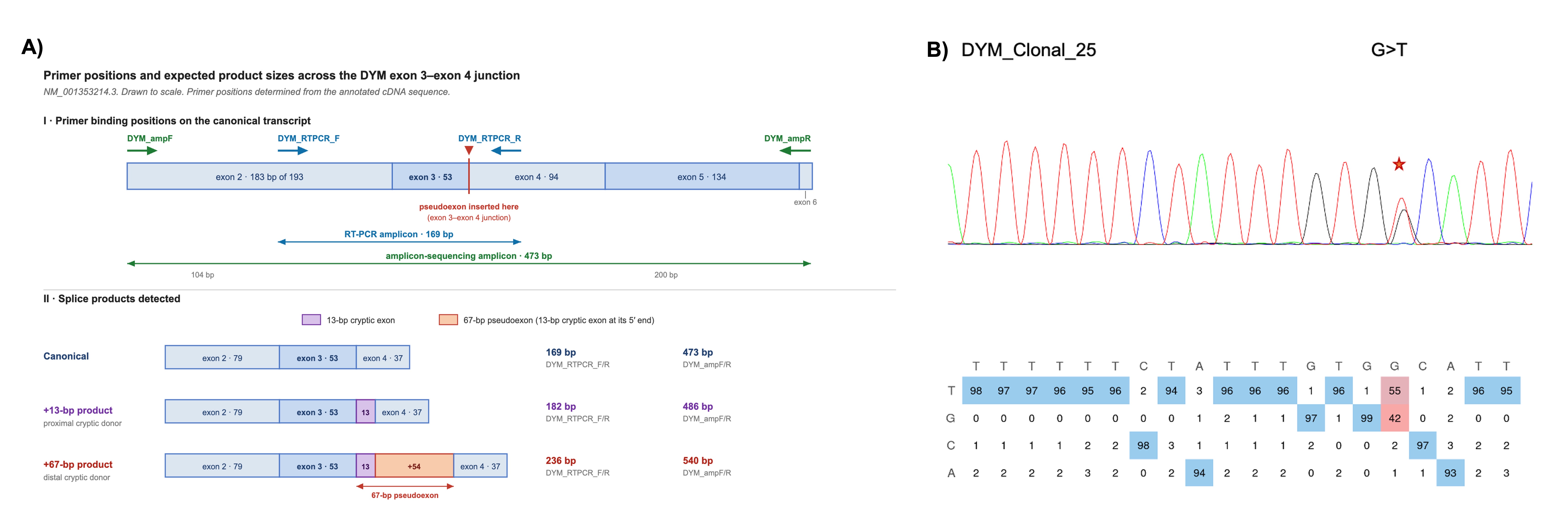

### Supplementary Figure 3

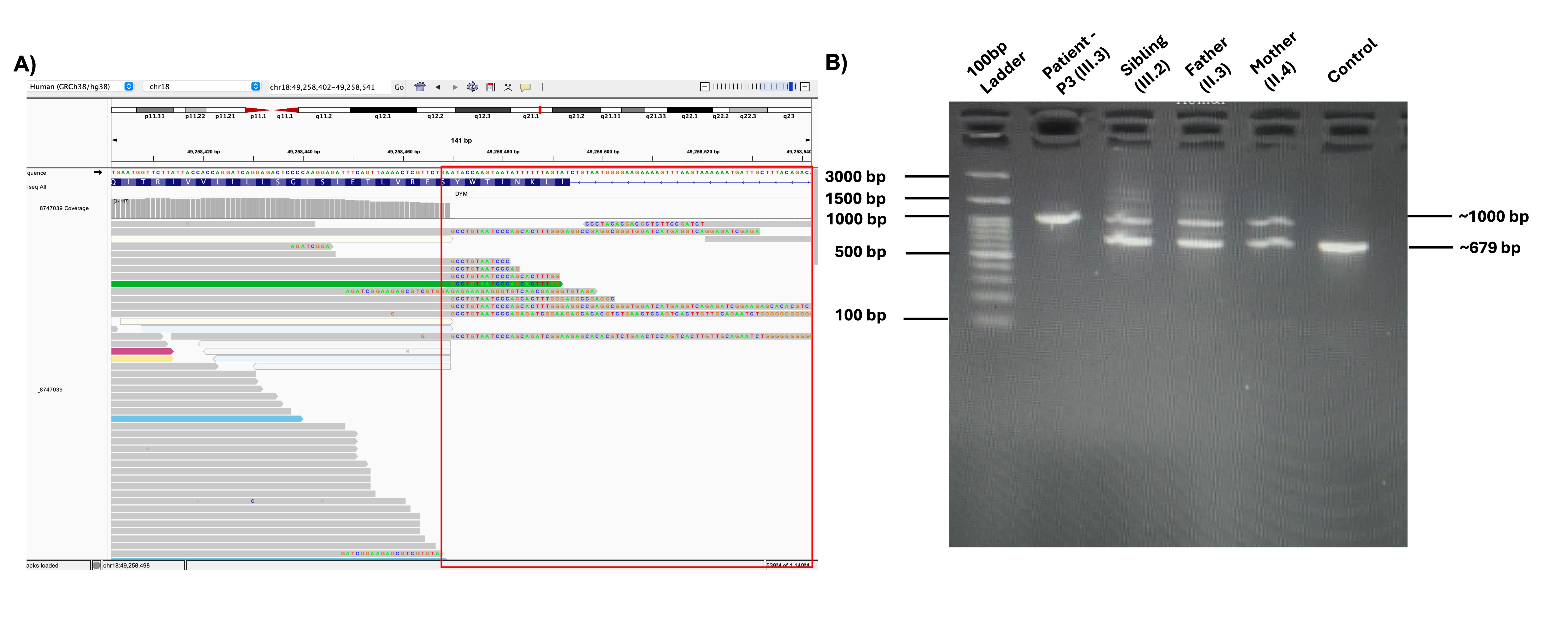

### Supplementary Figure 4

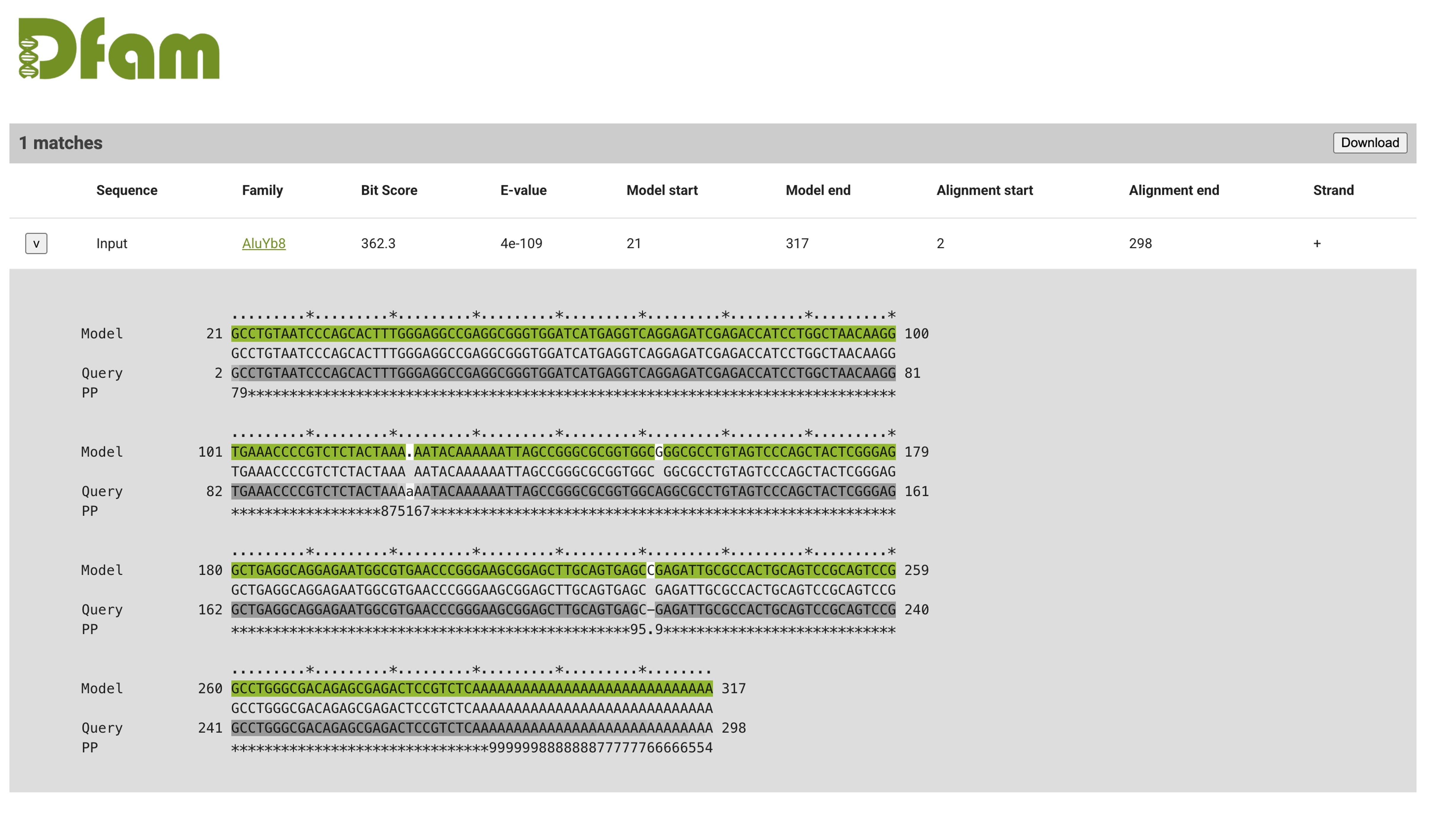
